# Causal Inference via Electronic Health Records in the National Clinical Cohort Collaborative: Challenges and Solutions in Long COVID Research

**DOI:** 10.1101/2025.06.06.25329168

**Authors:** Zachary Butzin-Dozier, Yunwen Ji, Lin-Chiun Wang, A. Jerrod Anzalone, Eric Hurwitz, Rena C. Patel, Mark J. van der Laan, John M. Colford, Alan E. Hubbard, the N3C Consortium

**Author notes:** Contributed equally to this work (Zachary Butzin-Dozier, Yunwen Ji). Members are listed at the end of the manuscript. **Correspondence:** Zachary Butzin-Dozier, PhD, MPH, Division of Biostatistics, School of Public Health, University of California, Berkeley, Berkeley Way West, 2121 Berkeley Way, Berkeley, CA 94720-7360, (302)437-6262.

## Abstract

Observational analyses of electronic health record (EHR) data using databases such as the National Clinical Cohort Collaborative include unique challenges for researchers seeking causal inferences, particularly when evaluating subjectively-defined outcomes like Long COVID. We explore several challenges and describe potential solutions. 1. Lack of true negatives: Many diagnoses and conditions either have a positive indicator or a missing status, requiring investigators to carefully consider which patients are likely negative for this condition. 2. Differential monitoring: EHR data include nonrandom missingness driven by patients engaging with the healthcare system at different rates, which is often related to both the exposure and outcome of interest. 3. Bias: EHR data sources face many biases, but are particularly vulnerable to informative missingness, differential monitoring, and model misspecification. 4. Large sample size: High precision (i.e., narrow confidence intervals) paired with potential bias leads to a high risk of incorrectly rejecting the null hypothesis. 5. Defining index time: It is important that investigators deliberately define index time (i.e., *t*_*0*_, baseline) to ensure that they only adjust for baseline confounders and do not adjust for (or condition on) factors that are affected by the exposure of interest (i.e., colliders or mediators). 6. Parameter selection: Investigators should only select parameters that are supported by the data distribution. This manuscript provides an overview of these challenges and solutions, using both simulated data and real-world data, with the outcome of Long COVID as the running example.

## Background

Randomized controlled trials are often considered the gold standard when evaluating a treatment’s causal impact on a health outcome of interest.^1–3^ Many research questions are poorly suited to randomized controlled trials due to ethical constraints (assignment to treatment or withholding treatment would violate equipoise) or logistical and financial constraints (lack of time, funding, or other resources). Furthermore, randomized controlled trials may have limited external validity, as the highly controlled trial context (and the participating trial sample) may not reflect the real-world conditions. Long COVID is an outcome where observational analyses are particularly relevant. As Long COVID is a rare outcome with a long induction period, randomized trials would need to enroll a very large sample size and conduct long-term follow-up in order to be sufficiently powered, which would be very expensive and resource-intensive.

Investigators often seek to draw causal inferences from observational data to address research questions that face these obstacles. This approach, known as the Causal Roadmap or an emulated target trial, seeks to reconstruct a scenario similar to a randomized controlled trial through observational data.^2–5^ This method starts with the question, “What would our ideal study look like?” and then seeks to approximate that design as closely as possible given the available data.^2,6–9^ There are many ways to pursue this approach, although a similar set of causal considerations persists. In particular, the importance of temporality (time ordering) of events is vitally important, as participant inclusion should not be based on a future factor, and analyses should not adjust for any factor that is a downstream effect of the intervention of interest.

Within the pharmacoepidemiologic space, an active comparator new user design allows for the strongest counterfactual comparator when an individual is included (i.e., “enrolled”) at the time of first prescription to the study drug or an active comparator.^10,11^ This moment of inclusion, which defines baseline (*t*_0_), is crucial as it defines the exchangeable populations that should theoretically be equivalent at baseline (assuming that the treatment of interest and active comparator are therapeutically equivalent and also that prescription is not based on confounding factors, such as SES).

Epidemiologic and public health evaluations of electronic health records (EHR) seek to estimate causal parameters that indicate the impact of a treatment (or exposure) on an outcome. Common parameters of interest in these settings include the average treatment effect and causal risk ratio. Using Targeted Machine Learning Methods, we can flexibly evaluate parameters such as optimal treatment regimens to identify targeted treatments for each patient and estimate the counterfactual outcomes under these optimal treatment schedules or marginal structural models to estimate complex, potentially continuous, treatment impacts of interest.^3,4,12,13^

A recent systematic review found a scarcity of causal inference methods applied to observational health data.^14^ The authors noted that modern approaches in data science and machine learning, combined with causal inference, are rarely applied to observational health data settings, and that filling this research gap may lead to significant advances in our understanding of human health. In this study, using evaluations of Long COVID in the National COVID Cohort Collaborative (N3C) as running examples, we aim to outline (1) major challenges to causal inference via observational EHR data, (2) potential solutions to these challenges, and (3) contrasts in estimates (using both simulated and observed data) using various approaches.

## Challenges and Solutions in Long COVID Research

### Lack of true negatives

EHR data often includes binary indicators (positive versus negative) for patient health conditions. Some measurements are routinely collected during most healthcare interactions (e.g., height, weight, temperature, blood pressure). On the other hand, many medical conditions are only documented when a case is suspected (e.g., flu). If a participant is never suspected of having a condition, there may be no mention of that condition in their EHR (i.e., no “negative” indicator). Therefore, it may be difficult to determine true negatives, as a patient missing a positive indicator for a condition does not necessarily mean that they did not experience that condition during the time interval of interest. Statistical models must deliberately account for nonrandom missingness, which is often driven by heterogeneous healthcare utilization.^15^

An unideal but straightforward solution to this challenge is restricting the patient sample only to include patients who engage with the healthcare system sufficiently to observe the relevant exposure, outcome, and covariate information. While this approach will ensure that the sample has high healthcare utilization, it may also induce bias (i.e., selection bias, collider stratification bias). In many cases, healthcare utilization may be associated with the exposure of interest and the outcome of interest. Therefore, conditioning on this behavior may induce a spurious relationship between the exposure and outcome (e.g., by creating a collider [ref] that induces confounding that did not yet exist). A seemingly logical next step may be to ignore healthcare utilization entirely, as it may be a consequence of the exposure of interest. Unfortunately, as healthcare utilization represents the patients’ probability of observation (for both the exposure and outcome), ignoring this factor will allow its biasing effect to persist.

One approach that defines relevant statistical parameters, but does not necessitate reliance on simple mis-specified (regression) models, is defining a variable of interest as the difference in the outcome distribution (usually mean) under theoretical interventions, termed counterfactual outcomes. Investigators estimate the intervention’s impact under a (counterfactual) scenario where healthcare utilization is held constant (intervened upon). The intervention should incorporate subject area knowledge as well as specific knowledge of the data (e.g., empiric distribution of healthcare utilization) to ensure that it provides sufficient participant observation without violating practical positivity assumptions (e.g., intervening to a frequency of healthcare utilization that is rarely observed in the sample population) (see *Parameter selection* below). Since these so-called causal parameters can rarely be estimated directly (except perhaps in a clinical trial), one must translate these causal parameters by equating them to parameters that can be estimated from the observed data, with explicit assumptions. This is referred to as the identification of a causal parameter as a statistical estimand.

Here, we provide simulated (Figure 1) and empirical (Figure 2) examples of how different methods to account for heterogeneous healthcare utilization can impact our estimations. For details on the data-generating distribution for our simulation and our estimators, see Supplemental Material 1.

**Figure 1.**
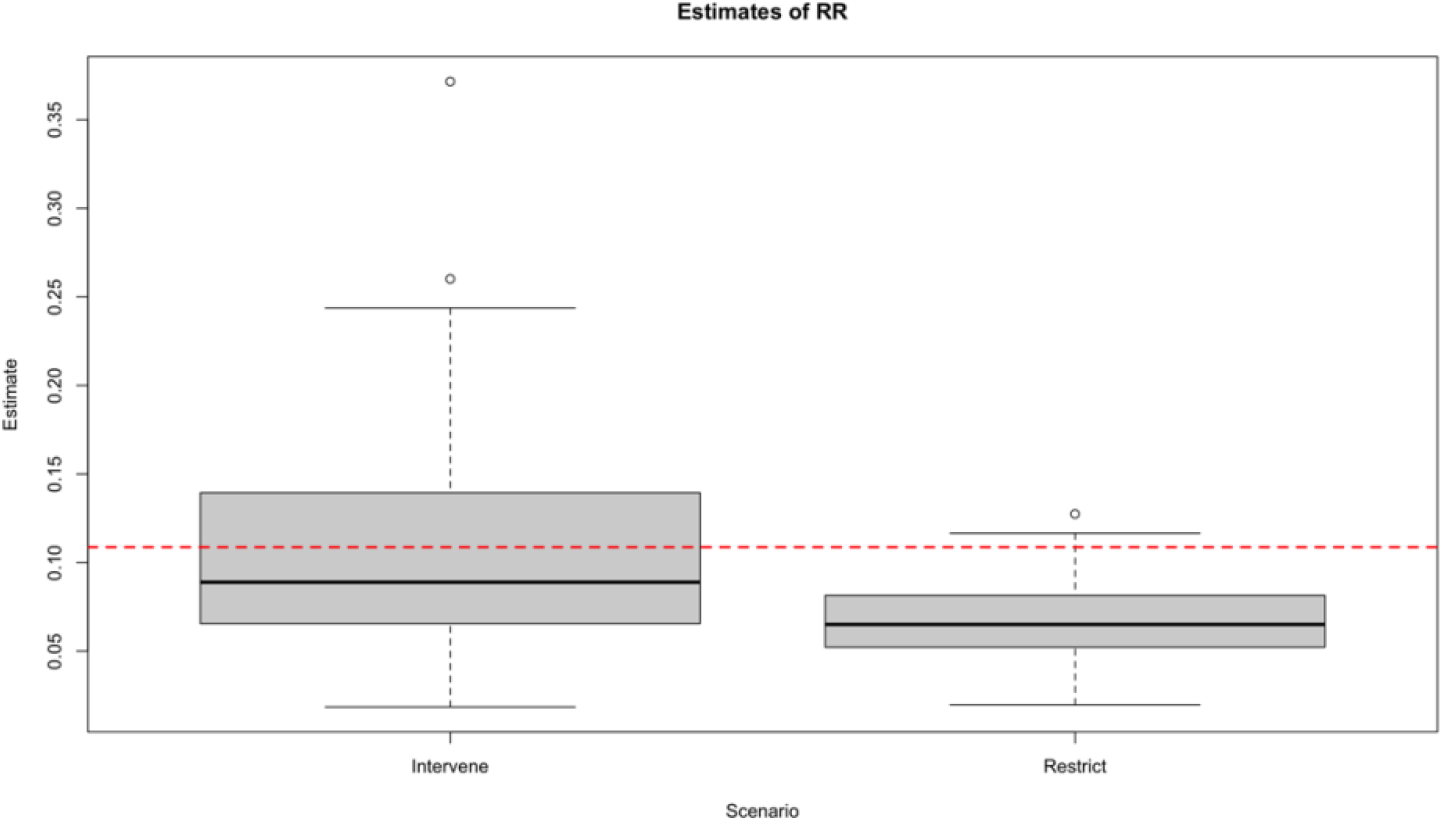
Simulation comparing intervention on observation versus restriction to observed individuals. “Intervene” refers to estimating the counterfactual outcome if all patients were observed (i.e., sufficient healthcare utilization), while “Restrict” refers to only estimating the outcome among patients who were observed. See Supplemental Material 1 for simulation details. RR: Risk ratio.

**Figure 2.**
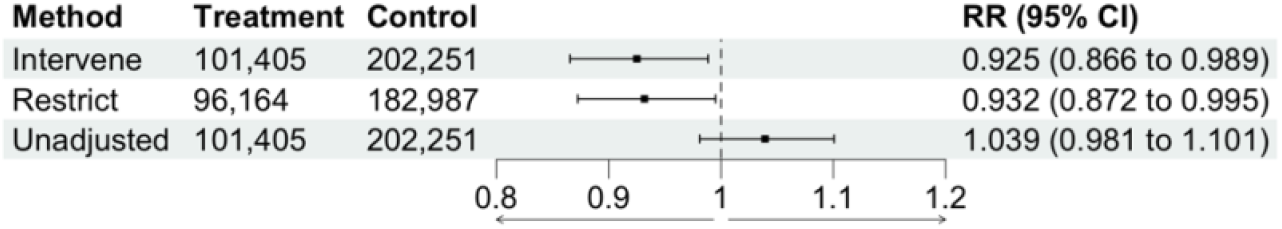
The relationship between SSRI use and Long COVID under different approaches to accounting for differential patient monitoring.^16^ “Intervene” refers to estimating the counterfactual outcome if all patients were observed at the desired level (i.e., at least one healthcare interaction in the year after acute COVID-19 diagnosis), while “Restrict” refers to only estimating the outcome among patients who were observed.

We observe *n* independent and identically distributed copies of *O* with a data structure *O* = (*W, A*, Δ, Δ*Y*) *∼ P*_0_ ∈ *M*, where *W* denotes patient covariates, *A* in (0, 1) the binary treatment variable, Δ the monitoring indicator, and *Y* the binary outcome observed only if Δ = 1. Our causal estimand is the risk ratio, Ψ = *E*_0_*Y*^1^/*E*_0_*Y*^0^. Two estimation methods are discussed in both examples. In the ‘restrict’ approach, analysis is limited to patients with Δ = 1 (i.e., those whose outcomes are observed). Under standard causal assumptions, the risk ratio is then identified as *E*_0_(*E*_0_(*Y* ∣ *A* = 1, *W*)/*E*_0_(*Y* ∣ *A* = 0, *W*)). In contrast, the intervention approach is to treat (*A*, Δ) as a joint intervention. The same assumptions identify the risk ratio as *E*_0_(*E*_0_(*Y* ∣ *A* = 1, Δ = 1, *W*)/*E*_0_(*Y* ∣ *A* = 0, Δ = 1, *W*)).

Our simulation shows that accounting for healthcare utilization by including it as a joint treatment provides a less biased (less confounded) measure of our risk ratio, demonstrated by the true value (red dotted line) falling near the center of our distribution. On the other hand, restricting our sample to only include patients with sufficient healthcare utilization induces significant bias, as demonstrated by the truth falling near the upper limit of our 95% confidence interval.

In a previously published study, we evaluated the relationship between selective serotonin reuptake inhibitor (SSRI) use during acute COVID-19 and subsequent risk of Long COVID.^16^ Similar to our simulation above, we intervened on patient observation to estimate the counterfactual outcome if all patients had been observed (i.e., all patients had a healthcare interaction in the 12 months following acute COVID-19). In this application, we found that ignoring healthcare utilization (unadjusted) led to a null estimate, while both using a restriction (exclusion) approach or an intervention (Observation intervention) approach led to observing a protective effect. Though the results show strong evidence of confounding from baseline confounders, there is little evidence that differential monitoring results in significant bias in this case. Of course, this is typically not known a priori. Therefore, the magnitude of bias exerted by applying a restriction vs. intervention approach to heterogeneous monitoring is often uncertain. Standard estimators of joint treatment–monitoring effects often break down when interval censoring creates near-positivity violations, a problem that is acute with fine time grids or when the monitoring process yields sparse data.^17^ An inverse-probability-weighting (IPW) approach based on the no-direct-effect (NDE) assumption, introduced by Neugebauer and colleagues^18^ and later compared with standard IPW by Kreif et al.,^19^ offers a remedy. The NDE-based IPW estimator relaxes the need for perfect adherence to the monitoring schedule; instead, it relies on a no-direct-effect assumption that the covariate trajectories would be identical under either monitoring regimen.

### Differential monitoring

Heterogeneous monitoring can be a significant source of bias in observational analyses of EHR data, as patient health information is only gathered via routine healthcare interactions. Therefore, EHR may have low sensitivity to detect short-term acute conditions and conditions defined by ambiguous or subjective criteria (e.g., Long COVID). EHR will detect more cases among patients who frequently interact with healthcare providers.^16,20^ Across medical systems and conditions, healthcare utilization rate is associated with the confounding factor of privilege (e.g., income level) and comorbidity burden.^20–23^

Censoring and loss to follow-up are often ambiguous in EHR data. Unlike a randomized trial or cohort study, EHR frequently lack information regarding patients’ loss to follow-up (e.g., relocating from a hospital catchment area). Therefore, it is often challenging or impossible to distinguish between patients who were censored from the study population versus patients who have not interacted with the healthcare system in the given time period.

### Bias

EHR often includes many sources of bias, which are usually challenging to identify or disentangle due to the researchers’ distance from data collection. This may consist of diagnostic bias, which can be driven by provider behavior or differential sensitivity of monitoring instruments.^24^ Geospatial trends may drive differences in diagnostic rates, whether from true geospatial trends in disease transmission, cultural norms around disease awareness and provider sensitivity, or heterogeneity in healthcare utilization. Heterogeneous documentation formats (e.g., OMOP, PCORNET)^25^ may also drive bias when conducting integrated analyses of multiple healthcare centers, as some documentation formats may be more sensitive than others for a given condition, and harmonization efforts may fail to capture all relevant data.

Given that EHR data are high-dimensional (many demographic variables and baseline measures of health per patient), model misspecification is of particular concern.^3,4,12^ These covariates allow investigators to adjust for various factors but require careful consideration of these relationships. Traditional parametric modeling approaches require the investigator to explicitly define the relationship between each covariate, the exposure, and the outcome, even when little theory exists to support such models, and assuming linearity for data with complex interactions can result in serious model bias. If the assumptions are violated, the model will be incorrectly specified, and the estimates will be biased. This bias is of particular concern in settings with a large sample size (see section “Large sample size and risk of Type 1 error”). Semiparametric estimation methods can reduce the risk of model misspecification by minimizing parametric assumptions. For example, a Super Learner ensemble machine learning approach includes a library of candidate algorithms rather than a single algorithm. It estimates the optimal combination of these algorithms to fit the data.^26,27^ The flexibility of this approach avoids model misspecification and bias, and does so implicitly as a function of sample size (as it goes up, more flexible models will be chosen because smaller non-linearities become important for prediction).

Generalizability and selection bias are particular concerns for studies involving EHR data. As mentioned previously, EHR will disproportionately represent highly privileged patients with a high comorbidity burden who frequently interact with the healthcare system. This is an obstacle to generalizability, or external validity, as estimates among these patients may not reflect impacts in the target population.

Investigators should critically evaluate the composition of their study population and how it may differ from their target population. The sample characteristics should be reported, and authors should highlight how differences between the sample and target population may impact generalizability. On the other hand, investigators can use a range of statistical techniques to address this source of bias, such as restriction, weighting (e.g., inverse probability of treatment weighting), resampling, or doubly-robust methods such as targeted maximum likelihood estimation.^2,9,12,12,13,28,29^

### Large sample size

The large sample size of EHR datasets enables meaningful evaluations of various health outcomes, including rare outcomes that would be underpowered in smaller samples. Unfortunately, for researchers using parametric analyses, this increased precision of a large sample size increases the probability of an incorrect rejection of the null hypothesis (i.e., Type 1 Error). In these settings, even minor bias (due to factors like model misspecification), paired with high precision (i.e., narrow confidence interval), will ensure that the confidence interval does not include the null hypothesis (a potential Type 1 Error). This type of error is particularly pernicious in scientific inference, as it leads an investigator to believe a relationship is present (e.g., “The drug works!”) while there is no true causal relationship. Null hypothesis significance testing is anchored on the principle that investigators should err on the side of failing to reject the null without strong evidence, but large sample size parametric analyses (particularly in high-dimensional contexts) will result in small p-value with near certainty.

**Figure 3.**
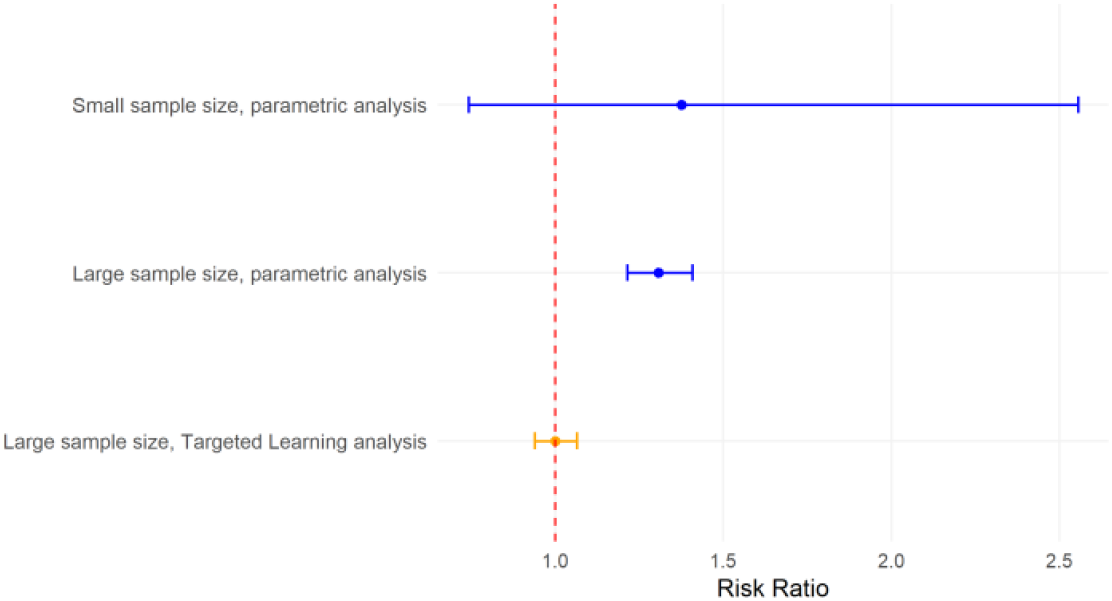
Risk ratio and 95% confidence interval in parametric versus Targeted Learning analyses, given a null relationship (i.e., risk ratio = 1.0) and moderate bias (e.g., model misspecification). See Supplemental Material 2 for simulation details.

A Targeted Machine Learning approach can address these issues in two ways. First, a Super Learner ensemble machine learning approach avoids model misspecification by creating a flexible combination of parametric and nonparametric algorithms (see “Bias”). Second, Targeted Maximum Likelihood Estimation is a bias-reduction step in which investigators can incorporate key information on the relationships between covariates (e.g., confounding, effect measure modification, and mediation). We have shown in simulations that, as a sample size gets very large in settings where the null hypothesis is true, Targeted Machine Learning estimates will converge on the null hypothesis, while parametric methods will converge excluding the null (Type 1 Error).^4^

### Defining index time (t_0_)

While modern epidemiologists have debated the necessity of many of the Bradford-Hill Criteria, temporality, or the need for exposure to precede outcome, has remained an essential tenet of causal inference.^30^ A fundamental yet often-overlooked challenge in making causal inferences via EHR is defining the patient index time (i.e., baseline, time of enrollment, *t*_0_). Disease-specific EHR sources, such as N3C, may implicitly define index time as the moment when a participant was diagnosed with the condition of interest. For N3C, this is the moment of COVID-19 diagnosis or the moment of a negative COVID-19 test for matched controls. While this may be an appropriate definition of *t*_0_ for studies evaluating the impact of COVID-19 on some future outcome (i.e., SARS-CoV-2 infection is the exposure of interest), this may be an inappropriate definition of *t*_0_ if one hopes to evaluate the impact of a previous exposure on risk of COVID-19 or a future outcome.

In our previous study, we sought to evaluate the impact of COVID-19 vaccination timing, relative to acute COVID-19, on subsequent Long COVID risk. For this study, we defined baseline time as the day of the first COVID-19 vaccination or booster dose between December 2021 and September 2022. We adjusted for patient information before baseline but did not adjust for factors occurring after baseline to avoid inducing bias. In addition, we included a sample of both SARS-CoV-2-positive and SARS-CoV-2-negative patients to avoid conditioning on COVID-19 status. As mentioned above, conditioning on COVID-19 status would induce bias in this setting, as acute SARS-CoV-2 infection and COVID-19 severity may mediate the relationship between COVID-19 vaccination and Long COVID. We intervened on acute COVID-19 infection timing to estimate the counterfactual impact of differing time between COVID-19 vaccination and COVID-19 infection on the subsequent risk of Long COVID.

In some settings, an active comparator new user design may not be possible. For example, in the study “SSRI use during acute COVID-19 and risk of Long COVID among patients with depression,” there was no therapeutically equivalent pharmaceutical intervention for depression that did not intervene (directly or indirectly) on serotonin.^16^ So, we defined exposure based on prevalent SSRI use during COVID-19 infection. This approach raised potential concerns regarding the exchangeability of exposed (SSRI users) and unexposed (SSRI nonusers) groups at the time of exposure assessment. To address this concern, we conducted a sensitivity analysis restricting our exposed group to only include new users of SSRIs within three months of COVID-19. We found a very similar relative risk in this subgroup analysis compared to our main analysis, which supported the fact that our observed effect estimate was not likely due to this bias.

**Figure 4.**
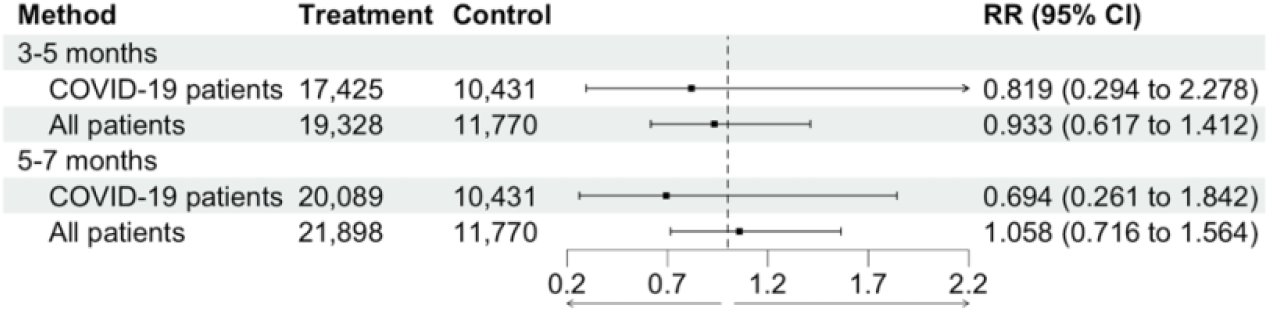
Comparison of effect estimates (risk ratios (RR)) for 12-month cumulative incidence of Long COVID after conditioning on diagnostic status (COVID-19 patients) versus including all patients.

Conditioning on diagnostic status may induce bias when evaluating an exposure that influences the diagnosis of this condition. Many EHR repositories sample patient records based on a given health outcome, such as N3C sampling patients diagnosed with COVID-19 (and sociodemographic-matched controls). This nonrandom sampling mechanism can lead to bias in several ways. If the exposure of interest influences the probability of SARS-CoV-2 infection or COVID-19 severity (e.g., COVID-19 vaccination), and the outcome is a consequence of COVID-19, then this sampling mechanism conditions on a mediator of the exposure-outcome relationship (as COVID-19 falls on the causal pathway between the exposure and outcome) and corresponding estimates will be biased. In addition, this sampling mechanism lends itself to defining the time of enrollment based on characteristics at acute COVID-19 (see *Defining index time* above), which is problematic if investigators hope to evaluate exposures that occur before COVID-19 and outcomes that occur after COVID-19.

### Parameter selection

Given the flexibility of Targeted Learning methods, it may be tempting to select innovative parameters that could rarely be estimated in a randomized setting. While this can be advantageous for exploring nuanced causal questions, it is crucial that even data-adaptive parameter selection is guided by the observed data distribution. Parameters should ideally be defined within a nonparametric or semiparametric model framework to avoid imposing restrictive parametric assumptions, which can introduce bias to the underlying causal structure. The investigator should ensure that the analysis approach aligns with the observed data and does not violate practical positivity assumptions. If treatment assignment is entirely determined by patient disease severity (i.e., patients with mild disease receive Treatment X, while patients with severe disease receive Treatment Y), it will be impossible to disentangle the relationships between treatment and the consequences of disease. In the context of Long COVID research, EHR analyses of treatments that are highly associated with COVID-19 severity, like Paxlovid, may face issues with positivity violations, as Paxlovid may only be prescribed to high-risk patients or patients with severe COVID-19, making these patients not comparable to patients not prescribed this medication. Parameter selection should be a compromise between (1) picking the most relevant causal parameter for the research question) and (2) ensuring that there is sufficient information in the observed data for estimation (i.e., avoiding positivity violations). This balance maintains the validity and interpretability of estimates.

### Conclusion

EHR data provide an excellent opportunity to investigate health relationships using large, high-dimensional datasets, but investigators must be careful to avoid common pitfalls. Long COVID is a subjectively defined outcome with a long induction period, making it highly vulnerable to censoring and missingness, which makes these considerations particularly relevant for investigators researching this condition.

## N3C Attribution

The analyses described in this manuscript were conducted with data or tools accessed through the NCATS N3C Data Enclave https://covid.cd2h.org and N3C Attribution & Publication Policy v 1.2-2020-08-25b supported by NCATS Contract No. 75N95023D00001, Axle Informatics Subcontract: NCATS-P00438-B, and [insert additional funding agencies or sources and reference numbers as declared by the contributors in their form response above]. This research was possible because of the patients whose information is included within the data and the organizations (https://ncats.nih.gov/n3c/resources/data-contribution/data-transfer-agreement-signatories) and scientists who have contributed to the on-going development of this community resource [https://doi.org/10.1093/jamia/ocaa196].

## Disclaimer

The N3C Publication committee confirmed that this manuscript (MSID: 2480.817, RP-884840) is in accordance with N3C data use and attribution policies; however, this content is solely the responsibility of the authors and does not necessarily represent the official views of the National Institutes of Health or the N3C program.

## IRB

This study was approved by the UC Berkeley Office for Protection of Human Subjects (2022-01-14980). The N3C data transfer to NCATS is performed under a Johns Hopkins University Reliance Protocol # IRB00249128 or individual site agreements with NIH. The N3C Data Enclave is managed under the authority of the NIH; information can be found at https://ncats.nih.gov/n3c/resources.

## Data availability

All analytic code and data are available in the N3C Enclave by request. Access to the N3C Data Enclave is managed by NCATS (https://ncats.nih.gov/research/research-activities/n3c/resources/data-access). Interested researchers must first complete a data use agreement, and next a data use request, in order to access the N3C Data Enclave. Once access is granted, the N3C data use committee must review and approve all use of data and the publication committee must approve all publications involving N3C data.

## Individual Acknowledgements For Core Contributors

We gratefully acknowledge the following core contributors to N3C:

Adam B. Wilcox, Adam M. Lee, Alexis Graves, Alfred (Jerrod) Anzalone, Amin Manna, Amit Saha, Amy Olex, Andrea Zhou, Andrew E. Williams, Andrew M. Southerland, Andrew T. Girvin, Anita Walden, Anjali Sharathkumar, Benjamin Amor, Benjamin Bates, Brian Hendricks, Brijesh Patel, G. Caleb Alexander, Carolyn T. Bramante, Cavin Ward-Caviness, Charisse Madlock-Brown, Christine Suver, Christopher G. Chute, Christopher Dillon, Chunlei Wu, Clare Schmitt, Cliff Takemoto, Dan Housman, Davera Gabriel, David A. Eichmann, Diego Mazzotti, Donald E. Brown, Eilis Boudreau, Elaine L. Hill, Emily Carlson Marti, Emily R. Pfaff, Evan French, Farrukh M Koraishy, Federico Mariona, Fred Prior, George Sokos, Greg Martin, Harold P. Lehmann, Heidi Spratt, Hemalkumar B. Mehta, J.W. Awori Hayanga, Jami Pincavitch, Jaylyn Clark, Jeremy Richard Harper, Jessica Yasmine Islam, Jin Ge, Joel Gagnier, Johanna J. Loomba, John B. Buse, Jomol Mathew, Joni L. Rutter, Julie A. McMurry, Justin Guinney, Justin Starren, Karen Crowley, Katie Rebecca Bradwell, Kellie M. Walters, Ken Wilkins, Kenneth R. Gersing, Kenrick Cato, Kimberly Murray, Kristin Kostka, Lavance Northington, Lee Pyles, Lesley Cottrell, Lili M. Portilla, Mariam Deacy, Mark M. Bissell, Marshall Clark, Mary Emmett, Matvey B. Palchuk, Melissa A. Haendel, Meredith Adams, Meredith Temple-O’Connor, Michael G. Kurilla, Michele Morris, Nasia Safdar, Nicole Garbarini, Noha Sharafeldin, Ofer Sadan, Patricia A. Francis, Penny Wung Burgoon, Philip R.O. Payne, Randeep Jawa, Rebecca Erwin-Cohen, Rena C. Patel, Richard A. Moffitt, Richard L. Zhu, Rishikesan Kamaleswaran, Robert Hurley, Robert T. Miller, Saiju Pyarajan, Sam G. Michael, Samuel Bozzette, Sandeep K. Mallipattu, Satyanarayana Vedula, Scott Chapman, Shawn T. O’Neil, Soko Setoguchi, Stephanie S. Hong, Steven G. Johnson, Tellen D. Bennett, Tiffany J. Callahan, Umit Topaloglu, Valery Gordon, Vignesh Subbian, Warren A. Kibbe, Wenndy Hernandez, Will Beasley, Will Cooper, William Hillegass, Xiaohan Tanner Zhang. Details of contributions available at covid.cd2h.org/core-contributors

## Data Partners with Released Data

The following institutions whose data is released or pending:

Available: Advocate Health Care Network — UL1TR002389: The Institute for Translational Medicine (ITM) • Aurora Health Care Inc — UL1TR002373: Wisconsin Network For Health Research • Boston University Medical Campus — UL1TR001430: Boston University Clinical and Translational Science Institute • Brown University — U54GM115677: Advance Clinical Translational Research (Advance-CTR) • Carilion Clinic — UL1TR003015: iTHRIV Integrated Translational health Research Institute of Virginia • Case Western Reserve University — UL1TR002548: The Clinical & Translational Science Collaborative of Cleveland (CTSC) • Charleston Area Medical Center — U54GM104942: West Virginia Clinical and Translational Science Institute (WVCTSI) • Children’s Hospital Colorado — UL1TR002535: Colorado Clinical and Translational Sciences Institute • Columbia University Irving Medical Center — UL1TR001873: Irving Institute for Clinical and Translational Research • Dartmouth College — None (Voluntary) Duke University — UL1TR002553: Duke Clinical and Translational Science Institute • George Washington Children’s Research Institute — UL1TR001876: Clinical and Translational Science Institute at Children’s National (CTSA-CN) • George Washington University — UL1TR001876: Clinical and Translational Science Institute at Children’s National (CTSA-CN) • Harvard Medical School — UL1TR002541: Harvard Catalyst • Indiana University School of Medicine — UL1TR002529: Indiana Clinical and Translational Science Institute • Johns Hopkins University — UL1TR003098: Johns Hopkins Institute for Clinical and Translational Research • Louisiana Public Health Institute — None (Voluntary) • Loyola Medicine — Loyola University Medical Center • Loyola University Medical Center — UL1TR002389: The Institute for Translational Medicine (ITM) • Maine Medical Center — U54GM115516: Northern New England Clinical & Translational Research (NNE-CTR) Network • Mary Hitchcock Memorial Hospital & Dartmouth Hitchcock Clinic — None (Voluntary) • Massachusetts General Brigham — UL1TR002541: Harvard Catalyst • Mayo Clinic Rochester — UL1TR002377: Mayo Clinic Center for Clinical and Translational Science (CCaTS) • Medical University of South Carolina — UL1TR001450: South Carolina Clinical & Translational Research Institute (SCTR) • MITRE Corporation — None (Voluntary) • Montefiore Medical Center — UL1TR002556: Institute for Clinical and Translational Research at Einstein and Montefiore • Nemours — U54GM104941: Delaware CTR ACCEL Program • NorthShore University HealthSystem — UL1TR002389: The Institute for Translational Medicine (ITM) • Northwestern University at Chicago — UL1TR001422: Northwestern University Clinical and Translational Science Institute (NUCATS) • OCHIN — INV-018455: Bill and Melinda Gates Foundation grant to Sage Bionetworks • Oregon Health & Science University — UL1TR002369: Oregon Clinical and Translational Research Institute • Penn State Health Milton S. Hershey Medical Center — UL1TR002014: Penn State Clinical and Translational Science Institute • Rush University Medical Center — UL1TR002389: The Institute for Translational Medicine (ITM) •Rutgers, The State University of New Jersey — UL1TR003017: New Jersey Alliance for Clinical and Translational Science • Stony Brook University — U24TR002306 • The Alliance at the University of Puerto Rico, Medical Sciences Campus — U54GM133807: Hispanic Alliance for Clinical and Translational Research (The Alliance) • The Ohio State University — UL1TR002733: Center for Clinical and Translational Science • The State University of New York at Buffalo — UL1TR001412: Clinical and Translational Science Institute • The University of Chicago — UL1TR002389: The Institute for Translational Medicine (ITM) • The University of Iowa — UL1TR002537: Institute for Clinical and Translational Science • The University of Miami Leonard M. Miller School of Medicine — UL1TR002736: University of Miami Clinical and Translational Science Institute • The University of Michigan at Ann Arbor — UL1TR002240: Michigan Institute for Clinical and Health Research • The University of Texas Health Science Center at Houston — UL1TR003167: Center for Clinical and Translational Sciences (CCTS) • The University of Texas Medical Branch at Galveston — UL1TR001439: The Institute for Translational Sciences • The University of Utah — UL1TR002538: Uhealth Center for Clinical and Translational Science • Tufts Medical Center — UL1TR002544: Tufts Clinical and Translational Science Institute • Tulane University — UL1TR003096: Center for Clinical and Translational Science • The Queens Medical Center — None (Voluntary) • University Medical Center New Orleans — U54GM104940: Louisiana Clinical and Translational Science (LA CaTS) Center • University of Alabama at Birmingham — UL1TR003096: Center for Clinical and Translational Science • University of Arkansas for Medical Sciences — UL1TR003107: UAMS Translational Research Institute • University of Cincinnati — UL1TR001425: Center for Clinical and Translational Science and Training • University of Colorado Denver, Anschutz Medical Campus — UL1TR002535: Colorado Clinical and Translational Sciences Institute • University of Illinois at Chicago — UL1TR002003: UIC Center for Clinical and Translational Science • University of Kansas Medical Center — UL1TR002366: Frontiers: University of Kansas Clinical and Translational Science Institute • University of Kentucky — UL1TR001998: UK Center for Clinical and Translational Science • University of Massachusetts Medical School Worcester — UL1TR001453: The UMass Center for Clinical and Translational Science (UMCCTS) • University Medical Center of Southern Nevada — None (voluntary) • University of Minnesota — UL1TR002494: Clinical and Translational Science Institute • University of Mississippi Medical Center — U54GM115428: Mississippi Center for Clinical and Translational Research (CCTR) • University of Nebraska Medical Center — U54GM115458: Great Plains IDeA-Clinical & Translational Research • University of North Carolina at Chapel Hill — UL1TR002489: North Carolina Translational and Clinical Science Institute • University of Oklahoma Health Sciences Center — U54GM104938: Oklahoma Clinical and Translational Science Institute (OCTSI) • University of Pittsburgh — UL1TR001857: The Clinical and Translational Science Institute (CTSI) • University of Pennsylvania — UL1TR001878: Institute for Translational Medicine and Therapeutics • University of Rochester — UL1TR002001: UR Clinical & Translational Science Institute • University of Southern California — UL1TR001855: The Southern California Clinical and Translational Science Institute (SC CTSI) • University of Vermont — U54GM115516: Northern New England Clinical & Translational Research (NNE-CTR) Network • University of Virginia — UL1TR003015: iTHRIV Integrated Translational health Research Institute of Virginia •University of Washington — UL1TR002319: Institute of Translational Health Sciences • University of Wisconsin-Madison — UL1TR002373: UW Institute for Clinical and Translational Research • Vanderbilt University Medical Center — UL1TR002243: Vanderbilt Institute for Clinical and Translational Research • Virginia Commonwealth University — UL1TR002649: C. Kenneth and Dianne Wright Center for Clinical and Translational Research • Wake Forest University Health Sciences — UL1TR001420: Wake Forest Clinical and Translational Science Institute • Washington University in St. Louis — UL1TR002345: Institute of Clinical and Translational Sciences • Weill Medical College of Cornell University — UL1TR002384: Weill Cornell Medicine Clinical and Translational Science Center • West Virginia University — U54GM104942: West Virginia Clinical and Translational Science Institute (WVCTSI) Submitted: Icahn School of Medicine at Mount Sinai — UL1TR001433: ConduITS Institute for Translational Sciences • The University of Texas Health Science Center at Tyler — UL1TR003167: Center for Clinical and Translational Sciences (CCTS) • University of California, Davis — UL1TR001860: UCDavis Health Clinical and Translational Science Center • University of California, Irvine — UL1TR001414: The UC Irvine Institute for Clinical and Translational Science (ICTS) • University of California, Los Angeles — UL1TR001881: UCLA Clinical Translational Science Institute • University of California, San Diego — UL1TR001442: Altman Clinical and Translational Research Institute • University of California, San Francisco — UL1TR001872: UCSF Clinical and Translational Science Institute NYU Langone Health Clinical Science Core, Data Resource Core, and PASC Biorepository Core — OTA-21-015A: Post-Acute Sequelae of SARS-CoV-2 Infection Initiative (RECOVER) Pending: Arkansas Children’s Hospital — UL1TR003107: UAMS Translational Research Institute • Baylor College of Medicine — None (Voluntary) • Children’s Hospital of Philadelphia — UL1TR001878: Institute for Translational Medicine and Therapeutics • Cincinnati Children’s Hospital Medical Center — UL1TR001425: Center for Clinical and Translational Science and Training • Emory University — UL1TR002378: Georgia Clinical and Translational Science Alliance • HonorHealth — None (Voluntary) • Loyola University Chicago — UL1TR002389: The Institute for Translational Medicine (ITM) • Medical College of Wisconsin — UL1TR001436: Clinical and Translational Science Institute of Southeast Wisconsin • MedStar Health Research Institute — None (Voluntary) • Georgetown University — UL1TR001409: The Georgetown-Howard Universities Center for Clinical and Translational Science (GHUCCTS) • MetroHealth — None (Voluntary) • Montana State University — U54GM115371: American Indian/Alaska Native CTR • NYU Langone Medical Center — UL1TR001445: Langone Health’s Clinical and Translational Science Institute • Ochsner Medical Center — U54GM104940: Louisiana Clinical and Translational Science (LA CaTS) Center • Regenstrief Institute — UL1TR002529: Indiana Clinical and Translational Science Institute • Sanford Research — None (Voluntary) • Stanford University — UL1TR003142: Spectrum: The Stanford Center for Clinical and Translational Research and Education • The Rockefeller University — UL1TR001866: Center for Clinical and Translational Science • The Scripps Research Institute — UL1TR002550: Scripps Research Translational Institute • University of Florida — UL1TR001427: UF Clinical and Translational Science Institute • University of New Mexico Health Sciences Center — UL1TR001449: University of New Mexico Clinical and Translational Science Center • University of Texas Health Science Center at San Antonio — UL1TR002645: Institute for Integration of Medicine and Science • Yale New Haven Hospital — UL1TR001863: Yale Center for Clinical Investigation

## Authors statement

Authorship was determined using ICMJE recommendations.

ZB and YJ: Generated research question, drafted manuscript, and managed project timeline.

LW, JA, EH, RCP, JS, ML, JC, and AH: Provided oversight on study design and analysis plan, supported analysis, reviewed manuscript, and provided feedback.

## Inclusion and ethics statement

All co-authors and collaborators included in this manuscript have fulfilled the criteria for authorship.

