## Supplemental Materials for "Causal Inference via Electronic Health Records in the National Clinical Cohort Collaborative: Challenges and Solutions in Long COVID Research"

April 24, 2025

### 1 Supplemental Material 1. Intervention vs. restriction simulation settings

We simulate i.i.d. observations

$$O : (W, A, \Delta, \Delta Y) \sim P_0,$$

where $W = (W_1, W_2)$ are baseline covariates, A is a binary treatment, $\Delta$ is a monitoring indicator, and $Y$ is a binary outcome observed only when $\Delta = 1$. The data-generating distribution is

$$W_1, W_2 \sim N\left((0.5, 1), \begin{pmatrix} 2 & 1 \\ 1 & 1 \end{pmatrix}\right),$$

$$A \sim \text{Bernoulli}(p), p = \Phi\left(0.1 - 0.45W_1 + 0.15W_2\right)$$

$$\Delta \sim \text{Bernoulli}(q), q = \Phi\left(0.6 - W_1 - 1.05W_2 + 3A\right)$$

$$U.Y \sim N(0, 1)$$

$$Y \sim \text{Bernoulli}(r), r = \text{logit}^{\text{-1}}\left(-3 - 3A + W_1 - 2W_2 + W_2 A + U.Y\right)$$

$$Y^* = \begin{cases} \text{NA} & \Delta = 0 \\ Y & \Delta = 1 \end{cases}$$

Our causal estimand is the risk ratio:

$$RR = \frac{EY^1}{EY^0},$$

where $Y^a$ denotes the counterfactual outcome under $A = a$.

We compare the following methods:

1. **Intervention**: We use the full data, including individuals with unobserved outcomes. Under standard causal assumptions, our target parameter can be identified as:

$$E_{P_0}(E_{P_0}(Y \mid A = 1, W, \Delta = 1) - E_{P_0}(Y \mid A = 0, W, \Delta = 1)).$$

2. **Restriction**: We restrict subjects with $\Delta = 1$. The observed data are reduced to $O = (W, A, Y)$. In this subset, the target paramter can be identified as:

$$E_{P_0}(E_{P_0}(Y \mid A = 1, W) - E_{P_0}(Y \mid A = 0, W)).$$

We performed 100 Monte-Carlo iterations, each with a sample size of 10,000. For every iteration, we applied TMLE in both the intervention and restriction scenarios and recorded the resulting point estimate of the risk ratio. We also computed the oracle risk ratio directly from $P_0$ to serve as a benchmark. We compared the empirical distributions of the two estimators across these 100 replications.

##### 1.0.1 Results

In addition to the Figure presented in the main text, Table 1.0.1 summarizes the bias, root-mean-squared error (RMSE), and 95% coverage for both estimators.

| Metric | Intervention | Restriction |
|---|---|---|
| Bias | 0.01343811 | -0.04320996 |
| RMSE | 0.09523496 | 0.04782918 |
| Coverage | 0.8 | 0.66 |

Table 1: Bias, RMSE and coverage of the two methods compared to the oracle risk ratio

### 2 Supplemental Material 2. Bias and sample size simulation settings

We consider a point-treatment setting with a binary treatment and binary outcome, generated according to

$$O : (W, A, Y) \sim P_0,$$

where

$$\begin{aligned} W_1 &\sim U(0,1), \\ W_2 &\sim U(0,1), \\ A &\sim Bernoulli(p), \text{with } p = \text{logit}^{-1}\left(0.25W_1^3 + 0.5W_2\right) \\ Y &\sim Bernoulli(q), \text{with } q = \text{logit}^{-1}\left(-1 + 0.3W_1^3 + 0.6W_1W_2\right) \end{aligned}$$

We simulate small-sample data ( $n = 150$) and large-sample data ($n = 10,000$). Each setting undergoes 500 Monte Carlo iterations, producing point estimates of the risk ratio and corresponding confidence intervals. The true risk ratio in this setup is 1 (indicating no treatment effect).

We compare the following methods:

1. **Parametric Analysis** using a generalized linear model, fitted with the misspecified formula $Y \sim A + W1 + W2$ (omitting interaction terms).

2. **TMLE Analysis**, in which the model forms are left unspecified, and we use a flexible library of learners (GLM, GAM, and XGBoost).
